# COVID-19 Epidemic in Switzerland: Growth Prediction and Containment Strategy Using Artificial Intelligence and Big Data

**DOI:** 10.1101/2020.03.30.20047472

**Authors:** Marcello Marini, Ndaona Chokani, Reza S. Abhari

## Abstract

Using a previously developed agent-based artificial intelligence simulation platform (EnerPol) coupled with ‘Big Data,’ the evolution and containment of COVID-19 in Switzerland is examined. The EnerPol platform has been used in a broad range of case studies in different sectors in all of Europe, USA, Japan, South Korea and sub Saharan Africa over the last 10 years. In the present study, the entire Swiss population (8.57 million people), including cross-border commuters, and the entire Swiss public and private transport network that is simulated to assess transmission of the COVID-19 virus. The individual contacts within the population, and possible transmission pathways, are established from a simulation of daily activities that are calibrated with micro-census data. Various governmental interventions with regards to closures and social distancing are also implemented. The epidemiology of the COVID-19 virus is based on publicly available statistical data and adapted to Swiss demographics. The predictions estimate that between 22 February and 11 April 2020, there will be 720 deaths from 83’300 COVID-19 cases, and 73’300 will have recovered; our preliminary variability in these estimates is about 21% over the aforementioned period. In the absence of governmental intervention, 42.7% of the Swiss population would have been infected by 25 April 2020 compared to our prediction of a 1% infection over this time period, saving thousands of lives. It is argued that future scenarios regarding relaxation of the lockdown should be carefully simulated, as by 19 April 2020, there will still remain a substantial number of infected individuals, who could retrigger a second spread of COVID-19. Through the use of a digital tool, such as Enerpol, to evaluate in a data-driven manner the impacts of various policy scenarios, the most effective measures to mitigate a spread of COVID-19 can be devised while we await the deployment of large-scale vaccination for the population globally. By tailoring the spatio-temporal characteristics of the spread to match the capacity of local healthcare facilities, appropriate logistic needs can be determined, in order not to overwhelm the health care services across the country.

## Introduction

COVID-19 is a highly transmittable viral infection, which is caused by the severe acute respiratory syndrome Coronavirus 2 (SARS-CoV-2), and is characterized by rapid human-to-human transmission.^1^ The World Health Organization (WHO) declared the outbreak of COVID-19 a public health emergency of international concern (PHEIC) on 30 January 2020. A rapid escalation of COVID-19 cases has been observed globally, as COVID-19 has high transmission rates as well as a relatively long incubation period,^2,3^ during which carriers without obvious symptoms can re-transmit to others. There are currently neither antiviral drugs nor vaccines available for COVID-19. It is anticipated that within 12 to 18 months, vaccines could become to be available for the general public; also, it is plausible that vaccines could be used in an earlier experimental phase for public health workers. Thus, the current crisis has forced public authorities around the world to manage this outbreak by limiting social contacts and by the extensive use of protective clothing and disinfectants. Additionally, limitations on the movement of people and the closing of national and international transportation have been implemented. The primary goal of public authorities is to manage the outbreak so as to avoid overwhelming the health care system, in addition to buy time in order to bring online more capacity and resources that can handle the current and potential future peaks in the demand for emergency services. As such, there is an acute need to better understand the temporal growth of the outbreak (both symptomatic and asymptomatic), the geographic distribution of the outbreak, and the evolution of hot spots in the outbreak.

Switzerland is one of the countries that is most affected by the COVID-19 pandemic. On 16 March 2020, the Swiss government declared an “extraordinary situation” over COVID-19, promoting social distancing measures, instituting a ban on all private and public events, closing schools nationwide, and closing places such as restaurants, bars, sports and cultural spaces; only businesses that provide essential goods remain open. These measures are in force until 19 April 2020. Effective 21 March 2020, the government further limited gatherings, throughout the country, to no more than 5 people. The Swiss government did not explicitly prohibit the movement of people but provided the population very stern recommendations to follow these rules. Based on the anecdotal evidence of relatively law-abiding nature of most of the Swiss population, it is expected that an overwhelming majority will follow the measures, which would significantly reduce the spread of the pandemic.

In this study, a prognosis of the evolution of the disease is predicted using an existing agent-based, artificial intelligence, simulation platform.^4^ The entire 8.57 million population of Switzerland, including social adaptation and current government policy, are considered. The economic slow-down and the transition in the use of Switzerland’s public and private transportation system are also considered. The different daily activities of all individuals in the population, accounting for the contact patterns of individuals during activities and on public transportation vehicles, are simulated with a stochastic model and averaged over 15-minute time intervals. Our existing epidemic transmission model^4^ is calibrated using recently available data for COVID-19; this model accounts for pre-intervention as well as post-intervention transmission rate, incubation period, and social distancing on the scale of individual persons. Furthermore, calibrated probabilistic sub-models account for demographics, and the period needed for recovery that are required to handle the COVID-19 cases.

## Methodology

Our agent-based simulation framework, EnerPol, which accounts for 100% of the entire population of Switzerland, and that was applied to scenarios of influenza in Switzerland,^4^ is used. EnerPol is a holistic agent-based model with choice models, where the agents adapt their behaviour through artificial intelligence as part of the solution. A schematic of EnerPol modules that are used for the agent-based simulation of epidemic spread is shown in Figure 1.

**Figure 1:**
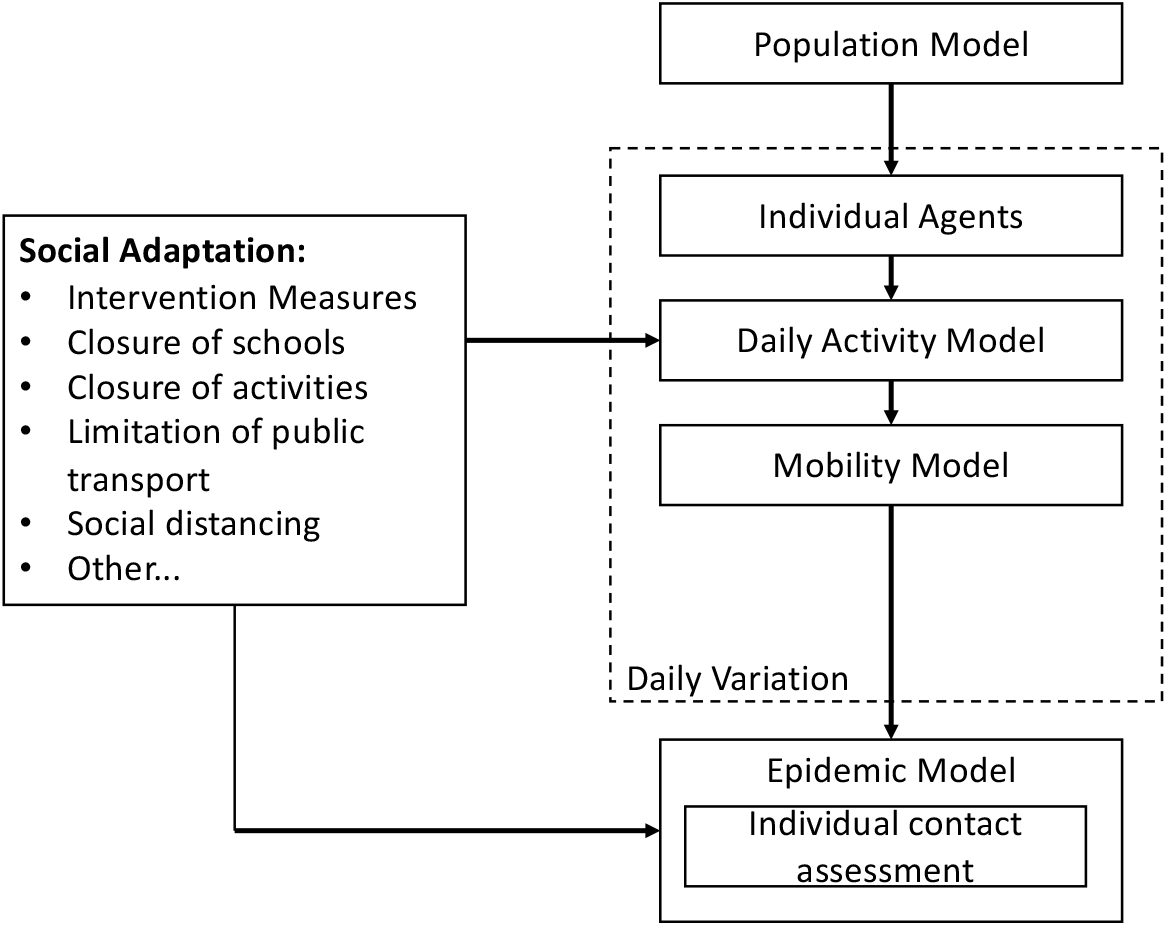
Schematic of the agent-based framework used for the simulation of epidemic spread.

The simulation framework is a fully parallelized time-marching algorithm optimized to run on GPUs such that a scenario covering 3 months can be completed within a few hours. The EnerPol platform has been used in a broad range of case studies in different sectors in countries throughout the world including; all of Europe, USA, Japan, South Korea and sub Saharan Africa. The present study, however, covers only the Swiss population, including cross-border commuters, and the entire Swiss public and private transport network. The synthetic population of 8.57 million individual agents, anonymized over samples of about 1’000 people, is generated from census data at the resolution of each of Switzerland’s 2’356 municipalities.^5^ Dwellings, workplaces, schools and other points of daily activity are derived from detailed federal registers, and each individual agent is linked, where relevant, to these locations. Thus, an activity-based demand for transportation is generated. As weather impacts possible daily activities, a mesoscale weather model that is integrated into the simulation framework is used to predict precipitation and ambient temperature. A detailed digital model of Switzerland’s road network (comprised of 1.1 million links and 0.5 million nodes), and public transit (30’000 stops and 20’000 routes) for trains, buses, tramways and other means of public transportation, is used to simulate Switzerland’s 3.5 million private vehicles and 1.7 million users of public transit with one-second resolution, and then aggregated over 15 minute time intervals.^6^ Therefore, contact patterns that could result in transmission of viral infections are modelled in detail with a spatial resolution of 1m. The likelihood of a viral infection and the subsequent transmission of the infection are modelled on an individual basis. The probability of being infected by the COVID-19 virus on contact with an infected person, is based on data from the spread of COVID-19 in South Korea,^7^ when the population was unaware of the state of the pandemic. Given the limited amount of data, a normal distribution of probability density function was assumed. The latent infection time for an asymptomatic person is specified to have a Poisson distribution, based on the best available fit to the South Korean data.^3^

Amongst the key factors in our modelling, are the exact date of the start of the spread in Switzerland and the seeding of the initial carriers within the population. Given the fact that in the early stage of the spread, testing in Switzerland was only performed for the 65+ demographic or for persons with chronic diseases, we matched the start of the spread for this demographic, and thereby estimated a mean probability density function of 1.4 × 10^−6^ per 15 minutes of contact with a mean of 10 days incubation, of which 8 days are infective, prior to the individual knowing that he/she is symptomatic. The mortality rate due to COVID-19 in multiple countries, including South Korea and Italy, has been shown to be strongly coupled to pre-existing conditions, specifically hypertension and heart disease, diabetics and lung disease.^8,9^ Given that the profile of chronic diseases in the Swiss population differs slightly from that in the South Korean population,^10,11^ the mortality rates observed in the spread of COVID-19 in South Korea were linearly adjusted for the case of Switzerland.

By performing a large number of simulations and machine learning steps, the approximate initial start date of the infection was determined to be Saturday 22 February 2020, when the simulation is seeded with 46 cases, whose geographic locations are based on the reported occurrences of COVID-19 in Switzerland.^12^ This date and seeding best match the observed ramp up of the spread of COVID-19 in Switzerland; this date and seeding were kept constant for the baseline study reported here. The simulations are perfomed with 15 minute temporal resolution for a 90 day period in order to evaluate the evolution of the spread of COVID-19, and the rate of recovery throughout the country.

## Results and discussion

Figure 2 shows the predicted geographic distribution of COVID-19 cases, at the resolution of Switzerland’s 2’356 municipalities, on four dates of 14 March, 21 March, 28 March and 11 April 2020. It is estimated that by 11 April 2020, there will be 83’300 COVID-19 cases. Figure 3 shows the evolution of recoveries from COVID-19. By 11 April 2020, 71’300 individuals will have recovered. As expected, the COVID-19 cases are concentrated in Switzerland’s largest urban populations of Zurich, Bern, Basel, Geneva, and Lausanne. Nevertheless, various other hot spots around the country are also observed. Based on multiple simulations that we conducted to assess the sensitivity of the predictions to different parameters including the profile of the seed carriers, initially we estimate an variability of +/- 21% in these predictions. Given the current urgency regarding the spread of COVID-19, we report with this level of variability based on our statistically insufficient sample of simulations; nevertheless, we continue to perform additional simulations to narrow the uncertainty estimate, and shall report this in future publications.

**Figure 2:**
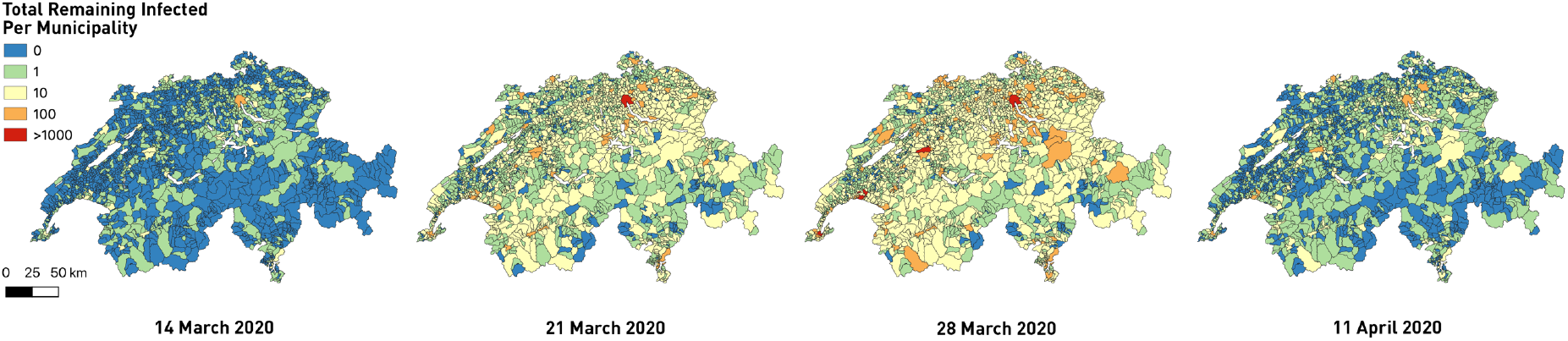
Geographic distribution of remaining COVID-19 cases (excluding recovered or dead) on 14 March, 21 March, 28 March and 11 April 2020.

**Figure 3:**
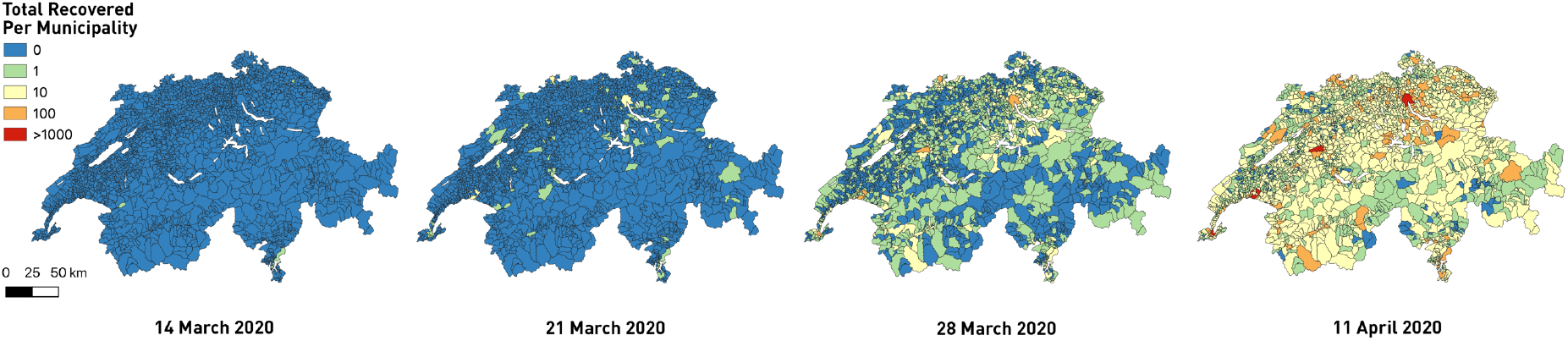
Geographic distribution of recovered COVID-19 cases on 14 March, 21 March, 28 March and 11 April 2020.

Due to a lack of available testing equipment, the Swiss authorities initially only tested the symptomatic individuals of the 65+ demographic, but then later added more testing for younger demographic groups. In Figure 4, we compare, on a daily basis, our predictions of the 65+ demographic COVID-19 cases to data from Switzerland’s Federal Office of Public Health (FOPH). It is not known to the authors whether, how, or over what time from the start of the pandemic, additional groups were also tested, which may partly explain the differences between our predictions and the data. Figure 5 compares the predicted and reported total numbers of death due to COVID-19. Our prediction estimates by 18 April 2020, a total of 720 deaths, of which 81% (586) are from the 65+ demographic.

**FIG 4:**
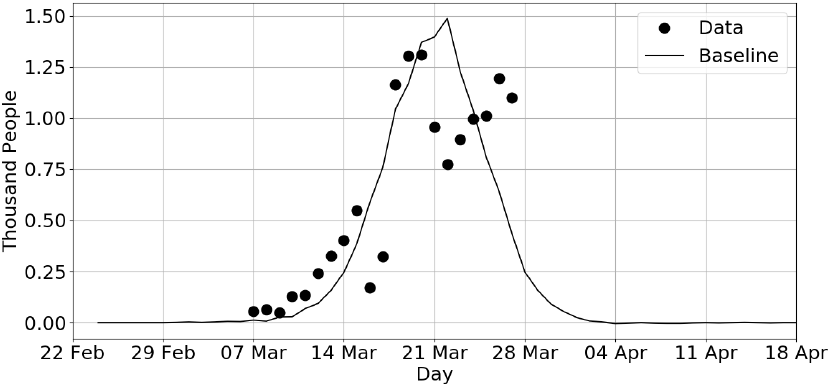
Comparison of predicted new infections to data for 65+ demographic for the period 22 Feb to 18 April 2020.

**FIG 5:**
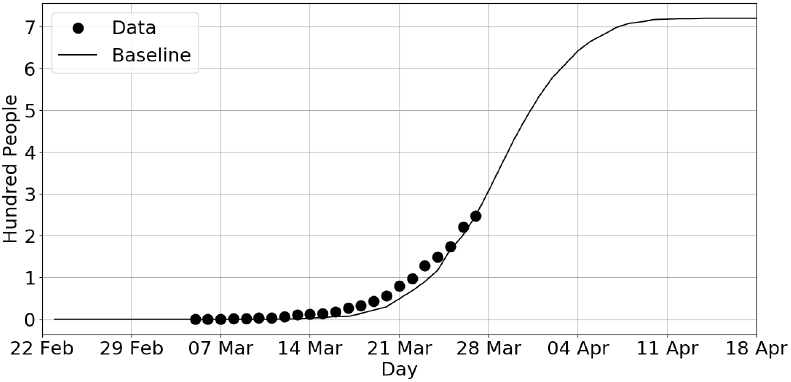
Comparison of predicted COVID-19 deaths to data for the period 22 February to 18 April 2020.

Figure 6 shows the temporal evolution in the total number of COVID-19 cases and recovered cases from 22 February to 18 April 2020. By the end of this period, the total number of COVID-19 infection cases is estimated to be 83’700. The recovered cases have a significant lag time compared to the total number of COVID-19 cases, due to the significant variability that is observed in COVID-19 patients. It should be noted that the COVID-19 cases include both symptomatic and asymptomatic cases, and as such the predicted number of COVID-19 cases is significantly higher than the officially reported confirmed COVID-19 cases which are a subset of the total number of cases.

**FIG 6:**
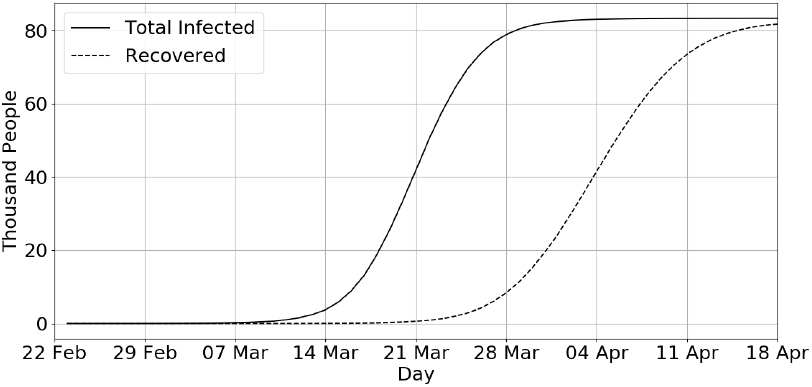
Predicted evolution of total infected and recovered COVID-19 cases for period 22 February to 18 April 2020.

The demographic distribution of COVID-19 infected individuals on 11 April 2020 is shown in Figure 8. It is evident that the 30 to 59 year age range constitue the majority of COVID-19 cases. The demographic distribution of deaths is shown in Figure 9. The vulnerability of those over 60 years is seen, and the extreme vulnerability of the 70-80 demographic is evident. Figure 10 shows the demographic distribution of mortality of in the Switzerland’s whole population of infected and uninfected persons. It can be seen that with the current government interventions, the mortality is below 1% for the entire population, but still a major cause for concern particularly for the elderly.

**FIG 7:**
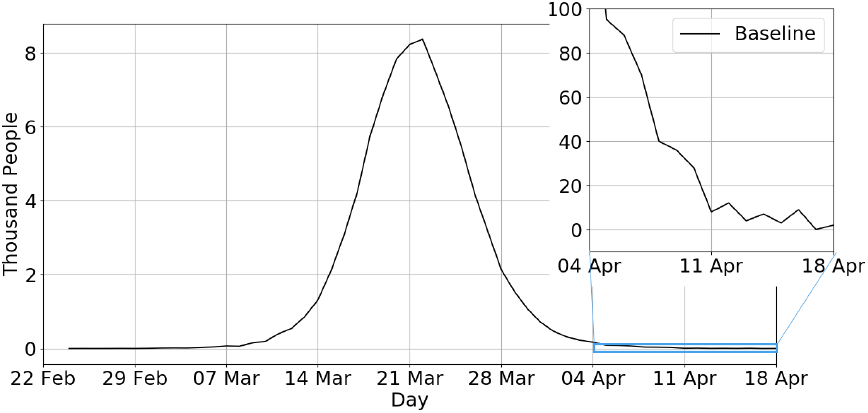
Predicted evolution of daily new infected COVID-19 cases for the period 22 February to 18 April 2020. Insert shows an expanded view for 4 April to 18 April 2020.

**FIG 8:**
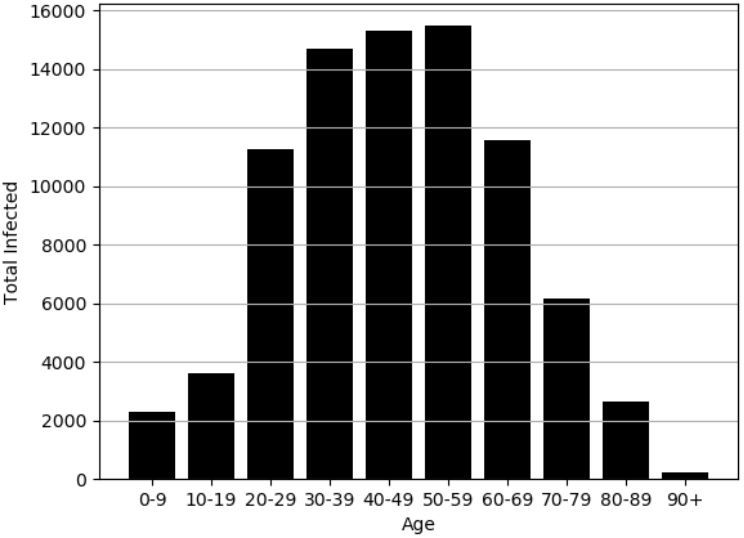
Demographic distribution of infected individuals on 11 April 2020.

**FIG 9:**
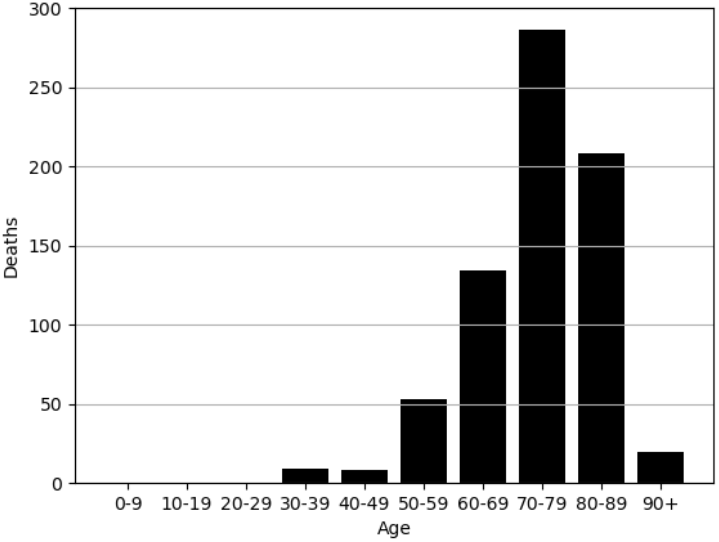
Demographic distribution of deaths from 22 February to 18 April 2020.

**FIG 10:**
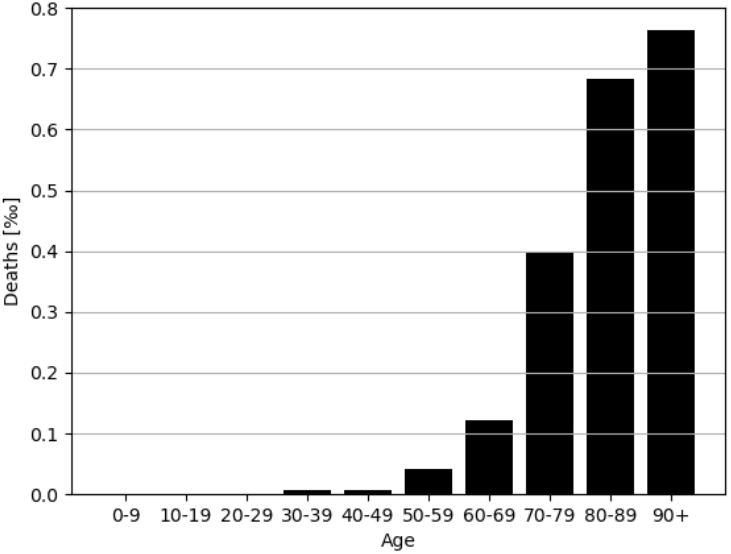
Demographic distribution of occurrence of death, for the whole population.

## Discussion and recommendations for future

Utilizing publicly available data, a holistic bottom-up agent-based simulation of the current COVID-19 pandemic in Switzerland is presented as a reference case. The initiation, growth and containment of the COVID-19 spread in presented, and quantified in terms of the infected (symptomatic and asymptomatic), recovered and deaths. Using the same simulation tool, it is shown that without social adaptation and governmental intervention, an explosive spread of the COVID-19 virus would have resulted in an infection of 42.7% of the entire population by 25 April 2020; on the otherhand the government’s timely intervention resulted in less than 1% of the population being infected for the examined time period. This shows that it is critical for goverments to step in, at an early stage to contain and manage pandemics and minimize mortality rates in the coming months. As restrictions become less prevelant, the infection rate and the associated mortality will undoubtedly increase.

As with any model simulation, there are uncertainties in our predictions. One of the key uncertainties is the exact behavioural profile of the first 50 or so seed carriers, whose contact behaviour results in variations in the initial phase of the growth of the pandemic. This illustrates the significance of early detection of new infected cases in all age groups, followed by contact-tracing of the individuals for the prior 8 to 10 days. Rapid and large scale testing would be crucial at this stage. The use of South Korean data in this study was very much linked to the appropriate reaction of the South Korean government to quickly test all suspected infected cases and not just the elderly. Other key improvements that are currently being developed is the need for the refining the future model fidelity by examining the exact trajectory of social behavior adaptation and modeling at a lower transmissivity rate, due to more disinfection of surfaces and contact avoidance.

Another issue of our existing model is that hospitals and health care facilities are not specifically differentiated due to the unavailability of reliable data. There seems to be some indication^13^ that, when health care facilities are overwhelmed by demand and have limited resources, these facilities themselves can become a hot zone of significant additional infections to staff and visitors. Once additional data become available, this limitation can be addressed in future work. As the first peak subsides, the role of imported new COVID-19 cases will again play a more significant role in the containment of COVID-19.

With large scale vaccination of the population some time away, the key question for the emergency management team would be on how to manage the pandemic, while minimizing the enormous economic and social impacts of isolation and the full-to-partial shutdown of the society including the transportation system and crossborder movement. Thus, in on-going work we are using scenario-based analysis to quantify the sensitivity of various interventions in order to better match the resulting demand for healthcare to the available local resources, in order to avoid overwhelming the system while waiting for the deployment of vaccination, which will hopefully come soon.

## Data Availability

All data used for validation and reference is publically available through the cited sources. Data generated by the developed models is not publicly available.

